# A cross-sectional survey of General Practitioners’ knowledge of the wait times for adolescent mental health specialists and services in Australia

**DOI:** 10.1101/2024.09.12.24312088

**Authors:** Bridianne O’Dea, Mirjana Subotic-Kerry, Thomas Borchard, Belinda Parker, Bojana Vilus, Frank Iorfino, Alexis E. Whitton, Ben Harris-Roxas, Tracey D. Wade, Madelaine K. de Valle, Nicholas Glozier, Jennifer Nicholas, Michelle Torok, Taylor A. Braund, Philip J. Batterham

**Author notes:** **Corresponding author:** Bridianne O’Dea Flinders University Institute for Mental Health and Wellbeing, Flinders University, Adelaide, South Australia, Australia, 5042.

## Abstract

**Background:** General Practitioners (GPs) play a key role in referring adolescents with depression and/or anxiety to mental health specialists and services, but their capacity to do so may be compromised by service wait times. It is unclear how GPs manage the mental healthcare of adolescents when the choice of treatment is not available. This study aimed to explore GPs’ self-reported referral practices to mental health specialists and services for adolescent depression and/or anxiety, as well as their perceived knowledge, acceptability, and impacts of the wait times for these.

**Methods:** A cross-sectional online survey of 192 GPs in Australia who self-identified as treating adolescents (12 to 17 years old) with depression and/or anxiety.

**Results:** GPs most frequently referred adolescents with depression and/or anxiety to psychologists. However, the mean estimated wait time for psychologists was 57.26 days (SD: 47.91, Mdn: 45.0, range: 5-365), which was four times the proposed acceptable wait time (M: 14.66 days, SD: 8.70). Nearly all GPs (81.8%) had increased their level of care for adolescents due to long waits but had limited training in and knowledge of strategies for effective self-management.

**Conclusions:** GPs in Australia lack information on the wait times for adolescent mental health specialists and services, despite frequent referrals. Greater knowledge of wait times, training in wait time approaches, and self-directed digital interventions may help to enhance the quality of primary care provided to adolescents.

## INTRODUCTION

General Practitioners (GPs) are the first source of professional help for many adolescents experiencing depression and anxiety and play a critical role in facilitating access to specialist treatment (Crouch et al., 2019; Lawrence et al., 2024; MacDonald et al., 2018). When treating mental illness, GPs are expected to follow clinical guidelines and escalate care as required. Australia’s clinical guidelines recommends specialists referrals for psychological therapy as first-line treatment for youth depression (Malhi et al., 2021) and anxiety (Andrews et al., 2018), with pharmacotherapies reserved for more severe cases with specialist supervision due to potential side effects. Other public mental health services (e.g. Headspace), community Child and Adolescent Mental Health Services (CAMHS) and hospital inpatient centres support primary care. However, psychological presentations to GPs have sharply increased over time, placing further strain on an already overburdened primary care system (Daneshmand et al., 2023; John et al., 2023; Russell et al., 2024; Villas-Boas et al., 2023). Mounting evidence suggests that GPs’ capacity to provide appropriate care and utilise referrals is increasingly limited by long wait times to mental health specialists and services (Dumesnil et al., 2018; Fleury et al., 2012; Paton et al., 2021; Petrie et al., 2021).

Despite no nationally agree-upon standards, up to 40% of Australians mental help-seekers waited ‘longer than acceptable’ for care (Mental Health Commission of New South Wales, 2024). On average, adolescents wait more than 100 days (Edbrooke-Childs & Deighton, 2020; Kowalewski et al., 2011; McNicholas et al., 2021; Punton et al., 2022; Subotic-Kerry et al., 2025) with similar or longer delays for psychiatrists and paediatricians globally (Edbrooke-Childs & Deighton, 2020; Kowalewski et al., 2011; Smith et al., 2018). Comparatively, systems in the UK, Norway and Canada follow wait time benchmarks of 4-6 weeks for psychological therapy. Delays to care represent a period of elevated clinical risk for adolescents and their GPs, as symptoms may worsen but treatment has not commenced. GPs often receive no information about expected wait times (Fleury et al., 2012) despite impacts on patient confidence and satisfaction (Bleustein et al., 2014). Faced with long waits, many adolescents return to their GP (Subotic-Kerry et al., 2025), placing further strain on practitioners who often lack the training, resources, and time to provide adequate interim support (De Silva et al., 2017). Despite these challenges, clinical guidelines offer little advice to GPs on how to proceed when first-line treatment is unavailable.

It is unclear how Australian GPs adapt to service shortages in youth mental health. A recent systematic review identified only 18 waiting list interventions, and none were initiated in primary care (Valentine et al., 2024). Some evidence suggests GPs have recommended lifestyle changes (e.g. exercise) and digital mental health tools (Keyes et al., 2023), scheduled follow-up appointments (Budd et al., 2021), delivered brief interventions through practice nurses (Koet et al., 2024) or medication (Ball et al., 2023). However short consultation times limit the extent of support that GPs can offer. Despite the known risks of delayed care, little is understood about how GPs manage adolescent mental health in the context of prolonged wait times.

### Aims of current study

This study explored GPs’ experiences of wait times for mental health specialists and services for adolescents with depression and/or anxiety in Australia. We examined their referral practices, awareness of wait times, perceptions of acceptable waits, and the impacts on GPs, patients, and treatment. Findings will inform how service availability affects GPs’ capacity to deliver timely, high-quality care to adolescents.

## METHOD

### Design

This study utilised a national cross-sectional online survey (co-designed with GPs) conducted in Australia between May 2022 to November 2022, at the end of the second wave of the COVID-19 pandemic. The sample size target was estimated on the proportion of GPs in Australia at the time of the survey (N=26, 599) who were likely to have mental health presentations from adolescents (overestimated to be 90%, given lack of available data), a 95% Confidence Level and margin of error of 5%. Based on this, the target sample was 379. The study was approved by the University of New South Wales Sydney Human Research Ethics Committee (HC220107).

### Participants, recruitment, and consent

Eligible participants were Australian GPs who self-reported treating adolescents (aged 12–17) with depression and/or anxiety and had verified registration with Allied Health Practitioner Regulation Agency (AHPRA) and the Royal Australian College of General Practitioners (RACGP) or the Australian College of Rural and Remote Medicine (ACRRM). Screening included registration verification, correct reporting of valid Medicare Benefits Schedule (MBS) item numbers and naming of their Primary Health Network (PHN). GPs were recruited via articles, GP magazines, paid and unpaid social media posts, GP conferences, PHN newsletters, and email lists from affiliated organisations. Advertisements linked to an online survey that contained study information and consent. Eligible participants received a $50 AUD voucher and were emailed study results.

### Measures

Survey items were adapted from prior studies (Fleury et al., 2012; Steele et al., 2010). See Supplementary Table S1 for full details.

GP Characteristics: Age, gender identity, work location (state/territory, metro vs rural/regional), years of experience, employment status (full-/part-time), and number of practices. GPs reported frequency of adolescent (12–17 years) consultations per week, proportion of adolescent patients, and proportion with depression/anxiety. They indicated if adolescent depression/anxiety presentations had increased, remained stable, or decreased over 12 months. Additional mental health training, specifically in adolescent mental health, and frequency of parent/guardian attendance during consultations were recorded.

Referral and Wait Times: Using 5-point Likert scales (never=1 to always=5), GPs rated how often they referred to mental health professionals/services or recommended other interventions. They estimated wait times (days) for subsidised/public services (psychologists, psychiatrists, headspace, CAMHS, inpatient centres), with an ‘unsure’ option. Acceptability of these wait times was rated on a 0–5 scale, and GPs indicated what they considered acceptable wait times in days. GPs reported whether they or their practice monitored wait times, and how helpful this knowledge was for treatment decisions. Using 5-point Likert scales (strongly disagree=1 to strongly agree=5), GPs rated agreement with statements on the impact of wait times on their duties and patient outcomes. They also indicated how often they referred adolescents elsewhere or increased their care due to long waits.

Care During Wait Times: GPs rated the importance of providing mental health strategies/resources to waiting adolescents (1=not at all to 5=extremely important). Frequency of recommending 13 listed interventions (e.g., sleep hygiene) was rated (never=1 to always=5), with free-text options for other interventions and suggestions on helpful supports during waits. GPs rated the quality of clinical support provided to adolescents and their parents/guardians (1=very poor to 5=excellent) and self-rated their knowledge of helpful strategies/resources during wait times (1=no knowledge to 5=superior knowledge).

### Data analyses

Qualtrics administered the survey. Invalid responses were determined by email/IP checks and response patterns (e.g., item responses, response speed). Suspected cases were reviewed by two researchers and discrepancies resolved by a third (see Supplementary Table S2). Analyses were conducted in SPSS. Participants’ postcodes were used to classify Geographic area was classified using postcodes and the Australian Government’s classification system. Wait times exceeding 12 months were excluded given referral expiry dates. For “select all that apply” items, percentages were based on total responses. To compare routine and wait-time care, mean scores were calculated and, where appropriate, combined into broader intervention types (e.g., diet and physical activity were grouped as lifestyle modifications). Free-text responses were analysed thematically by two independent raters, with disagreements resolved by a third. Associations between referral/recommendation frequencies and other variables were assessed using ordinal correlations (Spearman’s Rho), with a Bonferroni correction applied (*p* < .002). This was used to examine associations between GPs’ knowledge of wait times and background factors (e.g., experience, gender, weekly youth contact, adolescent patient load, mental health training) and referral frequency.

## RESULTS

### Sample characteristics

A total of 837 surveys were submitted, with 645 (77.1%) removed due to being invalid responses (*n=*523), ineligible (*n=*62), incomplete (*n*=55), or completed too quickly (*n*=7). The final sample consisted of 190 GPs. Most participants were female (71.6%, n=136), worked full-time (60.0%, n=114), and were based in a single practice (71.6%, n=136) in metropolitan areas (69.5%, n=132; see Supplementary Table S3 for state breakdown). GPs had a mean age of 42.38 years (SD = 10.25, range = 27–73) and 11.69 years of experience (SD = 9.93, range = 0.5–50). Two-thirds (66.8%, n=127) frequently treated adolescents (12–17 years), with most reporting adolescents made up a moderate (60.0%, n=114) to very high (12.1%, n=20) share of their caseload. The majority reported a moderate (46.3%, n=88) to high/very high (45.2%, n=86) prevalence of adolescent depression and/or anxiety, which had increased over the past year (80.0%, n=152). Parents were often or always present during consultations (57.9%, n=110). While 63.7% (n=121) had completed additional mental health training, fewer (28.9%, n=55) had specific training in youth mental health.

### GPs’ referrals and recommendations for adolescent depression and/or anxiety

As shown in Table 1, referral and recommendation frequency varied among GPs treating adolescent depression and/or anxiety. Most GPs (92.1%, *n*=175) “always” or “often” referred to psychologists (M = 4.24, SD = 0.67), while 39.5% (*n*=75) did so for headspace centres (M = 3.23, SD = 0.88). All GPs recommended ongoing GP care, lifestyle changes, and sleep hygiene to some extent, with moderate to strong positive correlations between these (*r* = 0.35–0.68, *p* < .001). Fewer than 1 in 10 GPs (7.9%, *n*=14) frequently prescribed medication. In free-text responses, some GPs recommended social/family/peer support (12.3%), mindfulness or meditation (8.2%), or further medical investigations (5.5%).

**Table 1.**
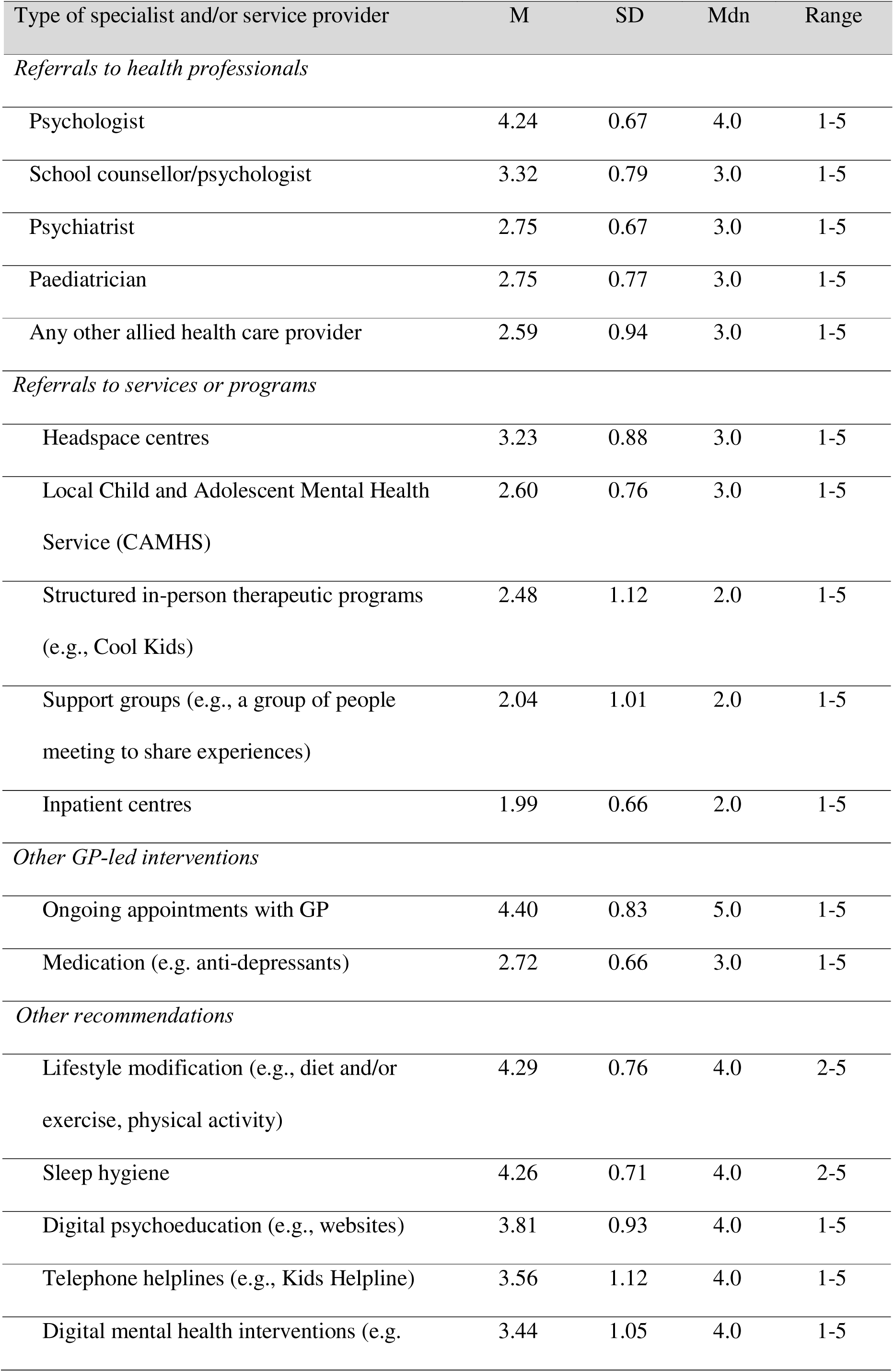
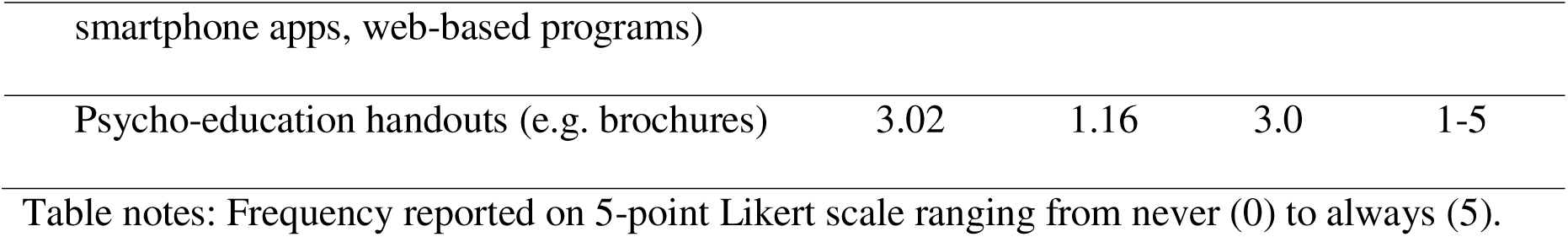
Frequency of GP referrals and/or recommendations to mental health specialists and services for adolescent depression and/or anxiety in Australia (N=190).

Several associations were observed between referral and recommendation types. Referrals to psychologists were linked only to ongoing care (*r* = .37, *p* < .001). Referrals to school counsellors were associated with headspace centres, ongoing care, and lifestyle changes (*r* = .25–.28, p < .001). Psychiatrist referrals correlated with paediatricians, CAMHS, and inpatient centres (*r* = .23–.37, *p* < .001). Referring to headspace was linked with CAMHS and allied health (*r* = .21–.23, *p* < .002). Medication recommendations were associated with CAMHS and inpatient referrals (*r* = .22–.24, *p* < .002), which may reflect variations in patient symptom severity.

### GPs’ knowledge of wait times for mental health specialists and services

Only 10 GPs actively tracked wait times (5.3%) and did so by contacting providers directly or by asking patients in follow up consultations. All but one participant (*n=*189, 99.5%) reported that up-to-date information on wait times would be helpful to some extent for informing their treatment decisions (M: 4.30, SD: 0.87, Mdn: 5.0, Range: 1 to 5).

As shown in Table 2, 27–37% of GPs were unsure about wait times for public mental health services (e.g., headspace, CAMHS, inpatient centres), despite referring to them, often for severe cases. No GP characteristics were associated with knowledge of wait times for psychologists, psychiatrists, headspace, or inpatient centres (*p* = .022–.707). Knowledge of CAMHS wait times was linked to a higher adolescent mental health caseload (*r* = –.24, *p* < .001). Wait time knowledge was generally unrelated to referral frequency, except for inpatient centres, where uncertainty was associated with higher referral rates (*r* = .24, *p* < .001).

**Table 2.**
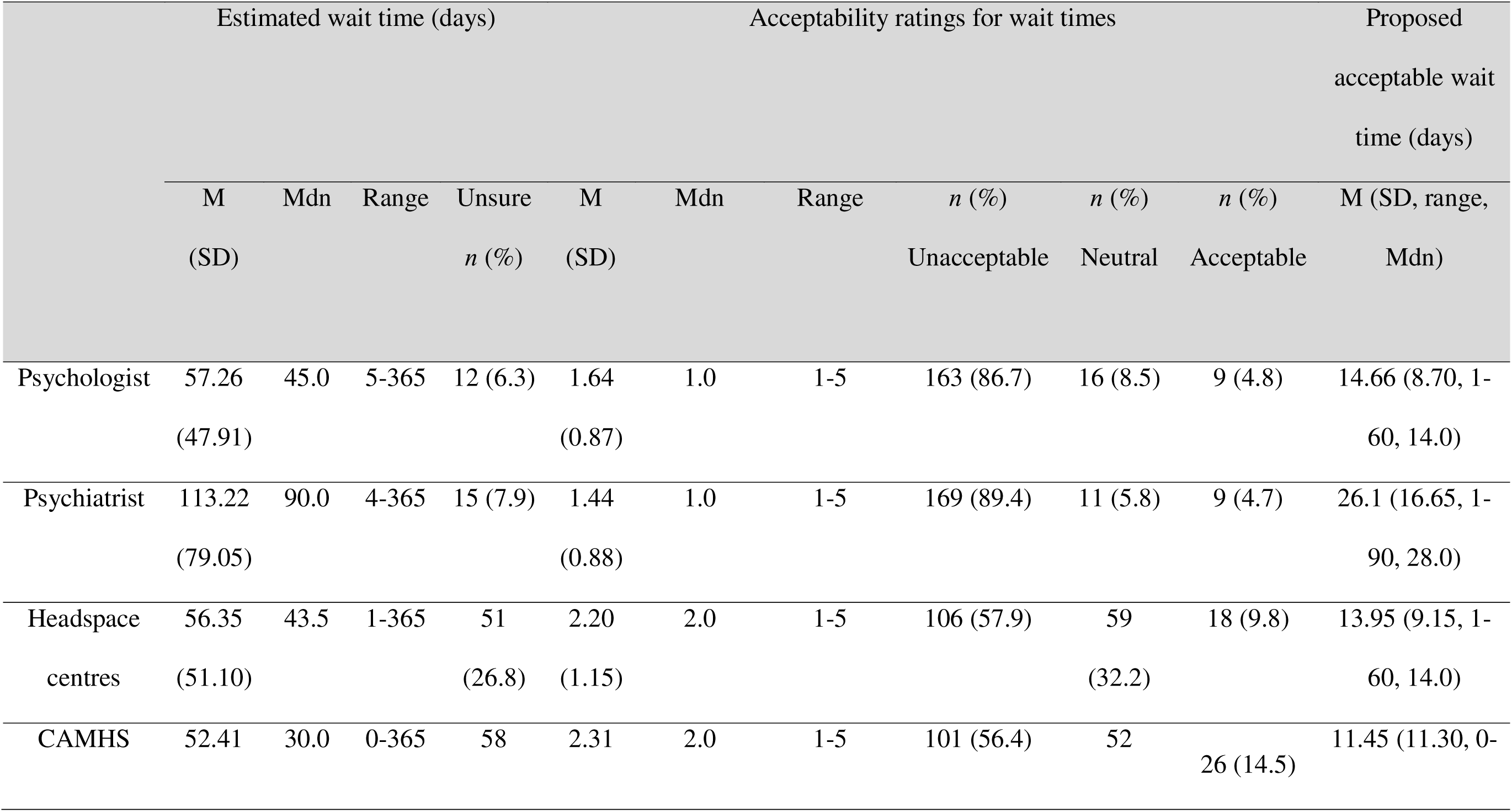
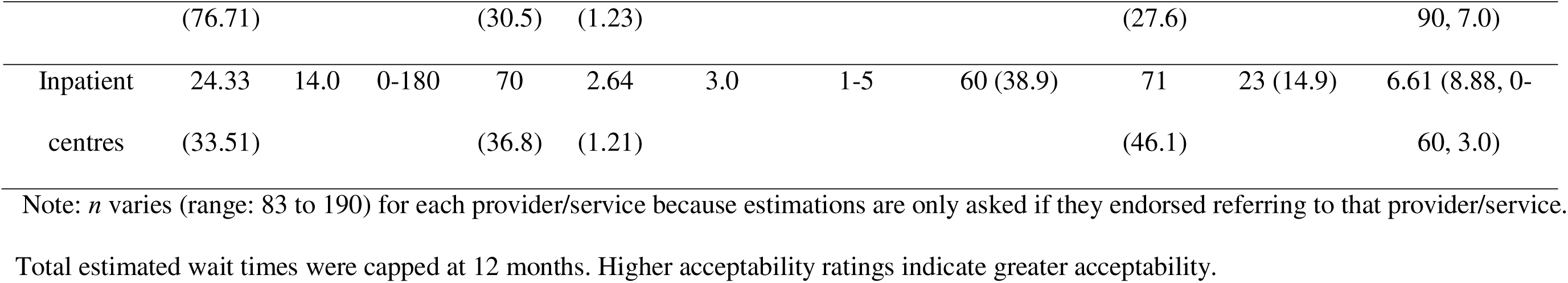
The mean estimated wait times for the frequently referred mental health treatment and services for adolescents and the ceptability of wait times among GPs in Australia (N=190).

GPs estimated the longest wait times for psychologists, headspace centres, and psychiatrists, ranging from 56 to 113 days (see Table 2). Metropolitan GPs reported significantly longer waits for psychiatrists (+39.48 days, *p* < .001) and CAMHS (+35.4 days, *p* < .002) than regional/rural GPs; no other geographic differences were found (*p* = .118–.378; see Table S4). Wait time estimates were unrelated to caseload factors (*p* = .080–.843). Most GPs deemed wait times for psychologists (86.7%) and psychiatrists (89.4%) unacceptable; over half said the same for headspace (57.9%) and CAMHS (56.4%). Across services, GPs considered acceptable wait times to range from 6.61 to 26.10 days—significantly shorter than their estimates (*p* < .001).

### The impact of wait times on GPs and their patients

Over two-thirds of GPs (70.0%, *n*=133) often or always referred elsewhere due to long wait times, and most (82.1%, *n*=156) reported increasing their level of care. As shown in Table 3, nearly all GPs agreed that long waits negatively affected their duty of care (96.3%), increased adolescent feelings of abandonment (94.2%), risk of dropout (95.7%), self-harm or suicide (90.5%), and reduced help-seeking (90.5%). In free-text responses, GPs also noted impacts on family/carer stress (10.5%), negative perceptions of the healthcare system (7.3%), and increased patient costs (5.2%).

**Table 3.**
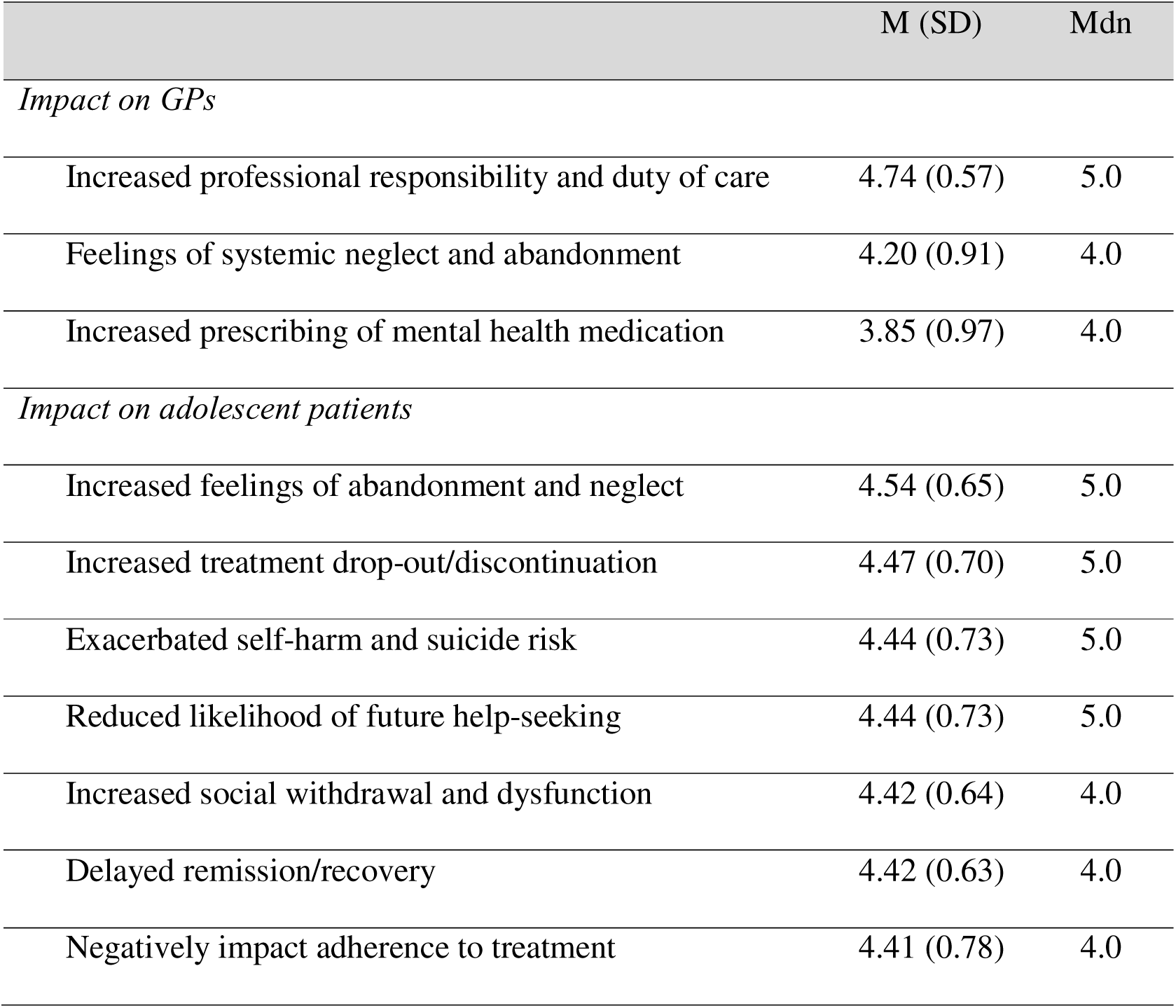
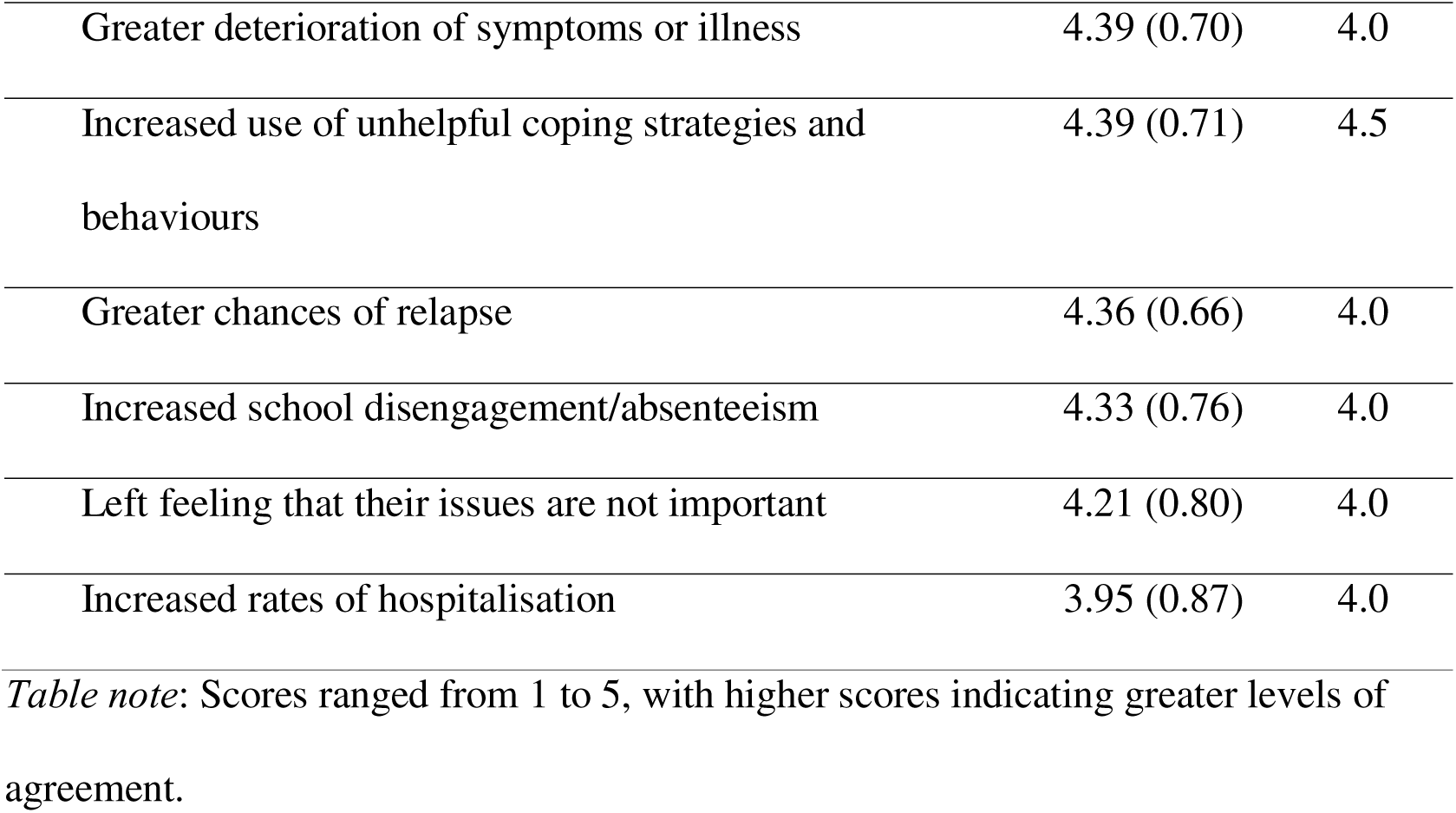
Level of agreement on the impacts of wait times on GPs and their adolescent.

GPs rated the quality of support provided to adolescents during wait times as fair (35.3%), good (50.5%), or excellent (6.3%) with a mean score of 3.53 (SD:0.79). Similar ratings were reported for care provided to parents/guardians (fair: 43.7%, good: 41.6%, excellent: 4.7%; M:3.39, SD:0.78). Nearly all GPs (95.8%) viewed it as very or extremely important to offer mental health strategies and resources during wait periods. However, most described their knowledge as minimal (8.4%), basic (37.9%), or adequate (48.4%), with only 5.3% reporting superior knowledge.

Commonly recommended interventions included ongoing GP appointments (88.9%), sleep hygiene (85.2%), exercise (84.7%), mindfulness (74.7%), and diet changes (66.8%). Over half also recommended mental health websites (67.8%), apps (55.2%), and web-based programs (50.5%). In free-text responses, GPs cited social support (21.1%) and relaxation activities (19.7%). As shown in Table 4, the frequency of interventions did not differ between routine care and wait-time care.

**Table 4.** A comparison of the frequency of interventions recommended by GPs in routine care for adolescent depression and/or anxiety with those recommended during the wait time (N=190).

| Type of intervention | Routine care<br>M (SD) | Wait time<br>M (SD) | Wait time<br>Mdn<br>(Range) |
| --- | --- | --- | --- |
| Ongoing appointments with GP | 4.40 (0.83) | 4.33 (0.70) | 4.0 (2-5) |
| Sleep hygiene | 4.26 (0.72) | 4.15 (0.72) | 4.0 (2-5) |
| Lifestyle modification (e.g., diet and/or<br>exercise, physical activity) | 4.29 (0.76) | 4.08 (0.81) | 4.0 (1-5) |
| Digital psychoeducation (e.g. mental health<br>websites) | 3.81 (0.93) | 3.92 (0.92) | 4.0 (1-5) |
| Digital mental health interventions (e.g.<br>smartphone apps, web-based programs) | 3.44 (0.93) | 3.47 (1.11) | 4.0 (1-5) |
| Telephone helplines (e.g., Kids Helpline) | 3.56 (1.12) | 3.56 (1.12) | 4.0 (1-5) |
| School counsellor/psychologist | 3.32 (0.79) | 3.49 (0.80) | 3.0 (1-5) |
| Psycho-education handouts (e.g. brochures) | 3.02 (1.16) | 3.30 (1.15) | 3.0 (1-5) |
| Medication prescribing (e.g. anti-depressants) | 2.72 (0.66) | 2.2 (0.75) | 3.0 (1-5) |

*Table note:* Scores ranged from 1 to 5, with higher scores indicating greater levels of agreement.

When describing ideal interventions for adolescents awaiting mental health treatment, GPs most frequently mentioned ongoing GP appointments (30.7%), digital interventions (28.1%), lifestyle changes (14.6%), multidisciplinary support (13.5%), and regular check-ins (13.0%).

## DISCUSSION

This study explored GPs’ self-reported referrals to mental health specialists and services for adolescent depression and/or anxiety in Australia, their estimates and acceptability of the wait times, and perceived impacts on treatment decisions. This study confirmed that GPs routinely referred adolescents with anxiety and/depression to specialist services with limited insight into anticipated wait times. GPs’ knowledge of wait times was not influenced by how frequently they referred patients, patient caseload, location, or training. Given that timely access to care is a key determinant of outcomes for adolescents, there is an urgent need to provide GPs with more precise estimates of expected wait times to enhance their referral decision-making and patient care. Overall, GPs felt that the wait times for almost all mental health specialists/services in Australia were unacceptable and should be less than one month. Information-sharing from psychologists who can accept new referrals should be prioritised, given they were the most common referral and often at capacity.

GPs agreed they were an important source of support in the wait time and had increased their involvement in care, consistent with reports by Australian adolescents (Subotic-Kerry et al., 2025). However, many interventions recommended by GPs during the wait time were the same interventions recommended in routine care. This suggests that GPs were not introducing supports specifically to address the unique needs of adolescents during the wait period. This may contribute to the sense of abandonment and neglect reported by youth (Subotic-Kerry et al., 2025). Digital resources warrant further investigation, as these can reduce symptoms without increasing service demand. Improving the digital literacy of GPs may increase the adoption of this approach to patient care during the wait time. Although lifestyle interventions are included in clinical guidelines for depression (Malhi et al., 2021), future research would benefit from clarifying whether GPs carefully assess the suitability of this self-management approach for adolescents. Physical activity is often the least preferred brief intervention among youth (Schley et al., 2019) with low-quality evidence to support its use (Oberste et al., 2020). The increased rates of disordered eating among adolescents (John et al., 2023) may also mean that some lifestyle modifications may be contraindicated. Screening tools may help GPs to tailor self-management recommendations while also helping to direct referrals more appropriately, ensuring patients are indeed waiting for the right provider.

GPs agreed that long wait times had a negative impact on their professional wellbeing. Specialised training in youth mental health and single session therapy (SST) approaches may increase GPs’ confidence and skills in managing the wait time. SSTs may be especially useful for GPs looking to improve the support provided to patients in follow-up appointments (Perreault et al., 2023). Family carers may be an underutilised support for GPs and adolescents during the wait time, given that many parents are present at appointments (Cronin et al., 2023). While parents have reported their own preferences for care during the wait time (Cunningham et al., 2013), future work should endeavour to bring GPs, adolescents, and their carers together to design a cooperative approach to wait time care that not only meets the psychological needs of adolescents but also strengthens family support and the capacity of GPs to provide high quality care.

### Limitations

Despite extensive recruitment efforts, our sample target was not reached. While larger than many prior studies, participants were younger and more likely to be female than national averages, aligning with data showing female GPs manage more youth mental health (Royal Australian College of General Practitioners, 2022). Referral patterns, wait time estimates, and acceptability ratings likely vary with case complexity, which was not assessed. Future studies should explore patient characteristics linked to increased risk during wait periods—such as developmental stage, socio-economic disadvantage, and parental separation. Qualitative methods (e.g. semi-structured interviews) could offer deeper insight into GP decision-making.

## Conclusion

Australian GPs have limited awareness of mental health service wait times but recognise that access to this information could improve referral and treatment decisions. While systemic efforts to reduce wait times are essential, GPs would benefit from training in strategies to support adolescents during these delays. Further research is needed to strengthen the supportive role GPs play in adolescent help-seeking.

## Statements and Declarations

### Author contributions

BOD conceived the project and led the funding acquisition with support from MSK, BP, AEW, BHR. BOD, BP, BHR, and TB led the development of the survey. BP, TB, MSK and EL provided research and operational support. MSK, TB, and BV analysed the data with statistical support from PJB. NG, JN, AEW, MT, TAB, and FI were chief investigators of some of the funding and provided scientific oversight. BOD and BV wrote the first draft of the manuscript with all authors providing feedback. All authors reviewed and approved the final manuscript.

### Data availability statement

A deidentified dataset is available by contacting the corresponding author.

